# Saving Lives with Pre-Financed Rules-Based Disaster Aid: Evidence from Mexico

**DOI:** 10.1101/2021.01.03.21249161

**Authors:** Alejandro del Valle

## Abstract

Developing economies are not disproportionately exposed to natural disasters, but they experience significantly more deaths. Exploiting a discontinuity in the eligibility rules to Mexico’s pre-financed indexed disaster fund (Fonden), I show that accelerated reconstruction of public infrastructure can fully reduce postdisaster excess mortality in the short-run and up to 75 percent two years after. I also find that Fonden’s impact is concentrated in areas with medical infrastructure and among conditions responsive to basic medical interventions. These findings suggest that Fonden operates by restoring access to health services after a disaster. Annual benefits from Fonden amount to 197 thousand life-years saved. (JEL: G22, H12, H84, Q54, I15, O18.)

---

In the last century, there have been 10,514 hydrometeorological events recorded and 8.3 million deaths that can be directly attributed to these events (CRED, 2020). The actual death toll is likely to be far larger as these records fail to account for indirect deaths, such as deaths attributable to the prolonged disruption of health and social services. While there are no global estimates of indirect deaths, evidence from Hurricane Maria in Puerto Rico indicates that total deaths could be more than 70 times larger (Kishore et al., 2018). Developing economies face the bulk of the burden because they experience most fatalities despite natural disasters not occurring disproportionately in these countries (Kahn, 2005; Strömberg, 2007).

A possible contributing factor to the larger death toll in developing economies is the bottlenecks in the disbursement of disaster relief, which results in delays in the restoration of critical services, including roads, safe water, and medical infrastructure. Two common sources of delays and leakage in the disbursement of disaster relief are reliance on postdisaster financing and lack of rules and administrative capacity to disburse these resources efficiently (Clarke and Dercon, 2016).

In this paper, I study Mexico’s Fund for Natural Disasters Fonden. This federal program is responsible for insuring public infrastructure and low-income housing against natural disasters. Fonden overcomes bottlenecks in the delivery of disaster relief by instituting and implementing a plan for disaster response. This plan has two key features. First, Fonden guarantees the availability of reconstruction funds before the occurrence of a disaster using a layered risk financing strategy. This strategy uses a protected federal budget allocation and Fonden reserves to pay for frequent small claims and relies on excess loss reinsurance and catastrophe bonds to pay for larger infrequent claims. Second, Fonden enforces a rules-based system for the disbursement of reconstruction funds. The rules define the process to verify a disaster’s occurrence (primarily using indexes), conduct damage assessments, disburse funds, and audit reconstruction projects.

To study whether Fonden saves lives in the aftermath of a disaster, I calculate for every municipality (the administrative unit below the state) the difference in the mortality rate before and after a disaster, at four-month intervals for two years. I then take advantage of the Fonden rules for disaster verification to derive causal estimates of the impact of Fonden on postdisaster excess deaths. Specifically, a municipality that experiences a hydrometeorological event (heavy rainfall, flooding, and tropical cyclones) is eligible for Fonden resources under the heavy rainfall rule if rainfall exceeds a predetermined threshold. This rule is the key of my research design because it allows me to estimate the impact of Fonden by comparing those municipalities that were barely eligible to those barely ineligible. Because municipalities can also become eligible to Fonden by meeting the tropical cyclone or flooding criteria (which I do not observe), I derive estimates of Fonden impact using a fuzzy regression discontinuity design.

The results fall into three categories. The first shows that Fonden can fully reduce post-disaster excess deaths and that this reduction, at least over the two years after the disaster, is mostly permanent. The second shows that Fonden reduces postdisaster mortality by restoring access to medical services. Specifically, I find that the impact of Fonden is driven by amenable conditions (conditions responsive to basic medical care) and that the Fonden led reduction in deaths from amenable conditions is only observed in municipalities where medical infrastructure was available before the disaster. I can also rule out that Fonden operates by reducing road-related deaths, deaths caused by inter-personal violence or suicide, or deaths caused by communicable conditions. The third reveals that consistent with the mechanism, adults 50 years or older are the demographic group that benefits disproportionately. Focusing on this group, I also document that the annual benefit from Fonden amounts to 197,000 life-years saved.

The paper is organized as follows. Section 1 summarizes the relevant institutional details of Fonden. Section 2 describes the data. Section 3 presents the identification strategy and results. Section 4 gives supporting evidence on the identification assumptions and robustness checks. Section 5 concludes by discussing the findings and their possible policy implications.

## 1 Disaster Response in Mexico

The Fund for Natural Disasters FONDEN was a program created by the federal government in 1996 to insure public infrastructure and low-income housing against geophysical and hydrometeorological perils. While Fonden insured several types of assets, the bulk of reconstruction expenditures corresponded to roads, hydraulic infrastructure, low-income housing, and educational and medical infrastructure. In December 2020, Fonden ceased operations when the government diverted resources from federal funds and trusts to pandemic relief programs. Since then, the responsibility of disaster response has devolved to state governments.

During its time in operation, Fonden reduced delays and improved the efficiency of reconstruction disbursements. Two design features made Fonden distinct from programs commonly used by other countries to fund and disburse disaster aid. First, Fonden relied on a financial plan to guarantee funds for a disaster of any size. The plan used the Fonden budget and reserves (0.4 percent of the federal budget on average roughly USD $800 million) to pay for frequently occurring small claims and relied on risk transfer instruments to pay for costly and infrequent claims. These instruments included a USD $400 million excess loss reinsurance policy on yearly payouts over USD $ 1 billion and several tropical cyclone and earthquake catastrophe bonds. In the unlikely event that catastrophe bonds failed to trigger and Fonden payouts exceed 1.4 billion, the program was designed to continue operating through an exceptional budget allocation from Mexico’s Oil Surplus Fund (World Bank, 2012).

The second design feature was a set of rules that defined the procedures for verifying the occurrence of a disaster, assessing damages, disbursing resources, and contracting and auditing reconstruction work. The Fonden process began with a request for disaster verification, which listed the affected municipalities. These requests were issued by the state government or by line ministries with affected assets. In the case of hydrometeorological events, which are the paper’s focus and account for as much as 93 percent of Fonden expenditures, the requests were routed to Conagua (the national water authority) for verification. This agency relied primarily on the heavy rainfall rule to corroborate the occurrence of a qualifying event. Specifically, this rule, introduced in 2004, established that a municipality is eligible for Fonden if daily rainfall, at any of the municipality’s representative weather stations, is greater or equal than the percentile 90 of maximum historic daily rainfall for the month in which the event took place. In addition to the heavy rainfall rule, municipalities can also become eligible for Fonden if they meet the flooding or tropical cyclone criteria. Specifically, Conagua verifies the occurrence of flooding if water has pooled in areas not normally submerged or if a body of water has overflown past its normal limits. Tropical cyclones are confirmed when Conagua observes sustained winds over 80 km/h.^1^

After the Conagua verification process is complete, the agency sends a report to SEGOB (the Ministry of the Interior) listing the municipalities that experienced a qualifying event. SEGOB additionally verifies that the disaster exceeds the local response capacity, which is almost always the case and issues a disaster declaration in the Federal Register (Official Journal of the Federal Government). The disaster declaration is a prerequisite for Fonden resources.

In the next step, a damage assessment committee, comprised of both federal and state representatives, visits the municipalities listed as eligible in the disaster declaration. The committee then issues a final damage report that includes itemized reconstruction costs, geocoded photographic evidence of damages, and proposed reconstruction improvements to reduce the likelihood of future damages. Fonden then verifies that there is no duplication of efforts, that resources are not used to repair preexisting damage, and that cost-sharing provisions have been enforced.^2^ The SHCP (the Ministry of Finance) is notified, and the disbursement of resources beings. In my sample, the disbursements begin on average within 75 days of the disaster, and most municipalities receive resources within 90 days. In 2009, Fonden further expedited the reconstruction of critical infrastructure by approving partial disbursements immediately after the Conagua verification.

In the last step, several federal agencies, such as the SCT (the Ministry of Communication and Transportation), assume the responsibility of designing and contracting the reconstruction work. While these agencies must provide Fonden with regular progress reports, they are allowed to follow their operating procedures and to hire third-party providers. The majority of reconstruction projects are completed within a year of fund disbursement.

## 2 Data

To study the impact of Fonden on excess deaths for up to two years after a disaster, I calculate the annualized mortality rate difference (AMRD) at four-month intervals. Specifically, I compute the difference in the mortality rate 4 months after a disaster with the same 4 month period’s mortality rate two years before the disaster. I then repeat this exercise using windows of 8, 12, 16, 20, and 24 months.

The mortality dataset is constructed in three steps. In the first step, I aggregate the 2000-2017 mortality records from Mexico’s national statistical institute (INEGI, 2017) to compute the count of deaths by municipality of residency and date of death (month-year). The counts include all certified deaths even if recorded with a five year lag or in a place different from the deceaseds municipality of residency.^3^ Certified deaths are likely to include all deaths. Mexico has met the World Health Organization (WHO) international reporting standards since the 1950s and routinely ranks in the top 20 of countries with high-quality mortality records (Mathers et al., 2005).

In the second step, for every municipality and month, I sum the number of deaths in the subsequent 4, 8, 12, 16, 20, and 24 months. I then use these counts to calculate crude annualized mortality *rates* by dividing the counts by the number of person-years at risk. In the cases where the population at risk is different from one year, I adjust the mortality rate denominator by multiplying the population by the fraction of the year that the population was at risk of dying. For example, this implies that the denominator is one-quarter of the municipal-year population in the case of the 4 months annualized mortality rate. The population data is drawn from the municipal year level projections from Mexico’s National Population Council (CONAPO, 2012, 2019). All rates are reported as deaths per 1,000 person-years.

In the third step, for every annualized mortality rate window (4, 8, 12, 16, 20, and 24 months), I calculate the annualized mortality rate *difference* (AMRD) by subtracting the corresponding annualized mortality rate (same window) from two years before. Note that by construction, the AMRD is computed from death counts that occur in the same calendar months. This feature of the calculation is important because deaths in Mexico are characterized by seasonal but stable trends.

Using an analogous three-step procedure, I also calculate AMRD’s that are cause-specific, age-specific, sex-specific, and age-sex adjusted (standardized). The cause-specific categories computed include amenable conditions (conditions responsive to basic medical interventions), unintentional injuries, self-harm and interpersonal violence, communicable, and noncommunicable conditions. The mapping between these categories and ICD-10 codes available in mortality records are given by Kruk et al. (2018) for amenable conditions and by GDB (2018) for all others. To calculate cause specific AMRD’s, I modify the first step and sum over the deaths attributable to the ICD-10 conditions that match each category. All other steps are as previously described. To calculate age-specific and sex-specific AMRD’s, I modify steps 1 and 2 so that the calculation only includes information from a given age or gender group. The age groups used are 0-14, 15-49, and 50 or older. To calculate the standardized AMRD, I modify the second step and calculate a weighted average of the previously derived age-sex-specific annualized mortality rates. The weights are given by the proportion of the age-sex group in the 2000 population.

Last, to perform falsification exercises, I modify step 3 and recalculate all the AMRD’s using only pre-disaster data. Specifically, I take the difference between the annualized mortality rates windows (4, 8, 12, 16, 20, and 24 months) from two years before and subtract the corresponding annualized mortality rate (same window) from four years before.

Next, I use information from federal disaster declarations that correspond to hydrometeorological events (heavy-rainfall, flooding, and tropical cyclones) to determine the sample of municipalities that requested Fonden resources and the subset that became eligible after verification. Specifically, I use the municipal month-year dataset assembled by Boudreau (2015), from disaster declarations published in Mexico’s Federal Registry between 2004 and 2012. I then replicate for this sample the Fonden verification process under the heavy rainfall rule. To this end, I use weather station identifiers to combine daily rainfall records from Conagua (2015*a*), and confidential records including the thresholds used for verification Conagua (2015*b*), and the mapping between municipalities and weather stations used for verification (Conagua, 2015*c*). Using the merged dataset, I then calculate the running variable by subtracting observed rainfall from the thresholds. Municipalities, where rainfall minus threshold is greater or equal to zero, are eligible for Fonden under the heavy rainfall rule. Because Fonden eligibility is triggered when the threshold is crossed at any weather station on any day, I report the running variable’s maximum in cases where municipalities have multiple weather stations or when events span multiple days in a month.

Using municipal and month-year identifiers, I then merge the dataset previously derived with the mortality dataset. To provide supporting evidence for the identifying assumption and to explore the channels through which Fonden operates, I extend this dataset with several other complementary municipal-year level datasets. These include data on health services infrastructure from the SSA (Ministry of Health), Fonden administrative records, and census and other datasets from INEGI (the National Institute of Statistics and Geography). A complete list of additional datasets and sources used can be found in table A1.

## 3 Results

### Fonden reduces postdisaster excess mortality

To identify the causal impact of Fonden on mortality, I exploit the discontinuous change in Fonden assignment at the heavy rainfall threshold and assume that the characteristics of municipalities that could affect mortality vary smoothly with the running variable (rainfall minus threshold). I provide supporting evidence for the identification assumptions in section 4. As previously mentioned, I use a fuzzy regression discontinuity (FRD) because a disaster occurrence can also be verified using the flooding or tropical cyclone criteria, and my weather dataset does not allow me to replicate the verification process using these criteria.

Figure 1 illustrates the FRD design panel A plots the fist stage and panel B the intention-to-treat. In each panel, the solid blue lines are fourth-order global polynomials fits. These lines are estimated separately on each side of the threshold and provide a smooth representation of the underlying conditional expectation. The red circles represent the local mean of the outcome over disjoint bins of the running variable. The green error bars are the 95 percent confidence intervals for the local means. Given the relative sparsity of observations far from the threshold, I use quantile space bins that are constructed to have roughly the same number of observations. The number of bins is chosen to minimize the integrated mean square error of the underlying regression function as described in Calonico, Cattaneo and Titiunik (2015).

**Figure 1:**
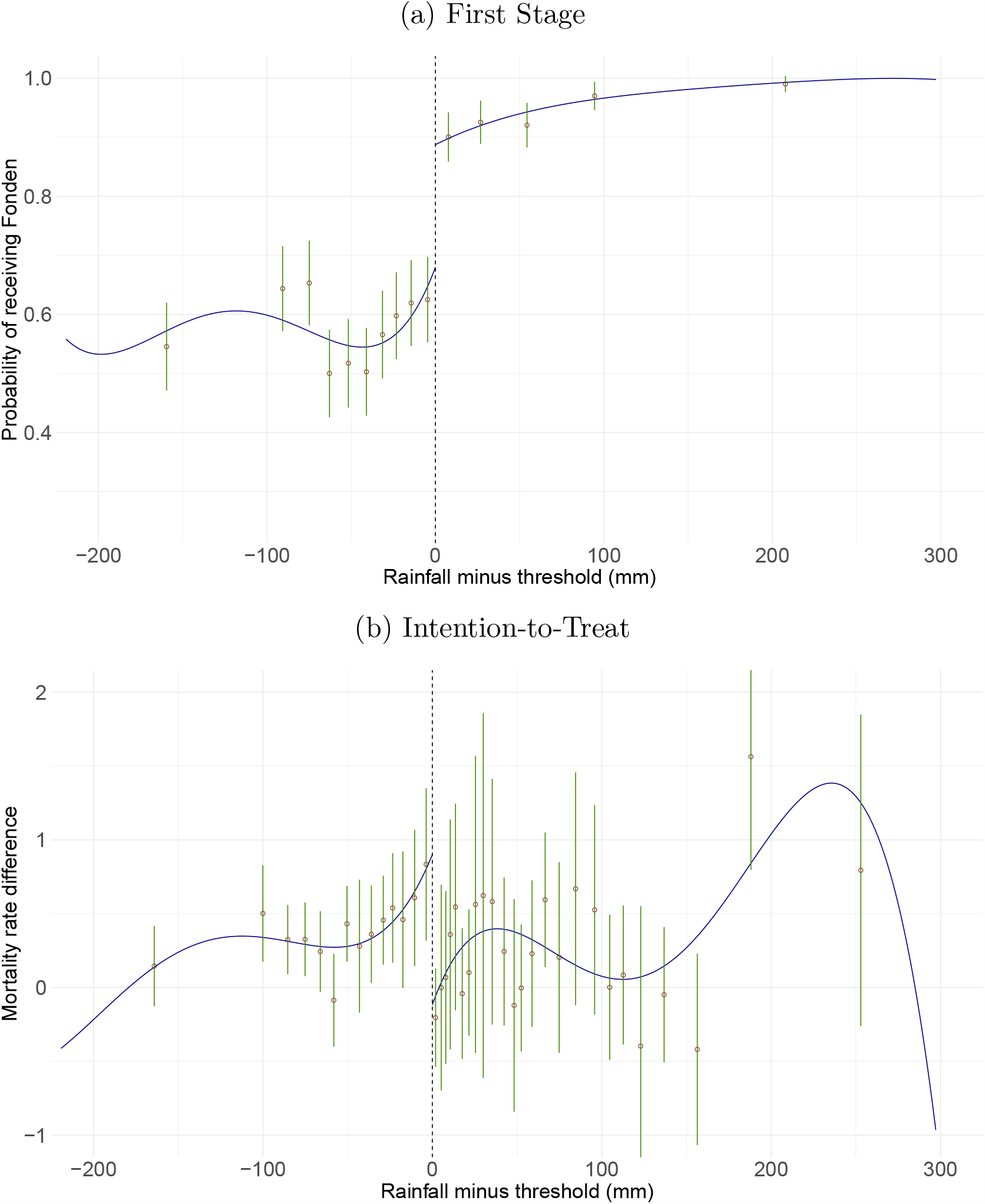
First Stage and Intention-to-Treat. Notes: Each graph plots the outcome (probability of receiving Fonden or 8 month annualized mortality rate difference) as a function of the running variable (rainfall minus threshold). In each graph, the support of the running variable has been partitioned into disjoint bins of roughly the same number of observations. The number of bins is selected to minimize the integrated mean square error of the underlying regression function, as described in Calonico, Cattaneo and Titiunik (2015). The circles plot the local mean of the outcome at the mid-point of each bin. The error bars are the 95% confidence intervals for the local means. The solid lines are fourth-order global polynomials fits (estimated separately from the raw data on each side of the threshold). Observations to the right of the vertical dashed line are eligible for Fonden.

In Panel A, the outcome is the probability of receiving Fonden. The figure reveals a clear first stage with the likelihood of receiving Fonden sharply increasing from 0.65 just below the threshold to 0.87 just above. This jump implies that the Fonden Local Average Treatment Effect (LATE) will be approximately 4.5 times larger than the Intention-to-Treat (ITT).

In Panel B the outcome is the 8-month AMRD, that is, the difference between the 8-month annualized mortality rate postdisaster and the same rate computed two years before the disaster. AMRD values greater than zero indicate that a municipality experienced excess deaths in the aftermath of a disaster. I use the 8-month AMRD for the figure because as shown in table 1 this is the window when Fonden leads to the most considerable reduction in excess mortality. The figure reveals a clear downward jump at the threshold and highlights that municipalities ineligible for Fonden (under the heavy rainfall rule) experience progressively larger increases in postdisaster excess mortality, with municipalities immediately to the left of the threshold experiencing roughly 0.79 excess deaths per 1,000 person-years. By comparison, municipalities eligible for Fonden, immediately to the left of the threshold, don’t experience any excess deaths.

**Table 1:** Evolution of Fonden Impact on all-cause AMRD.

|  | 4 months<br>(1) | 8 months<br>(2) | 12 months<br>(3) | 16 months<br>(4) | 20 months<br>(5) | 24 months<br>(6) |
| --- | --- | --- | --- | --- | --- | --- |
| Panel A. *First Stage* | 0.220 | 0.222 | 0.223 | 0.226 | 0.232 | 0.239 |
| *p*-value | $p < 0.01$ | $p < 0.01$ | $p < 0.01$ | $p < 0.01$ | $p < 0.01$ | $p < 0.01$ |
| CI 95 percent | [0.11,0.28] | [0.12,0.29] | [0.12,0.28] | [0.12,0.29] | [0.13,0.28] | [0.14,0.29] |
| Panel B. *Intention to Treat* | -0.502 | -0.816 | -0.491 | -0.412 | -0.334 | -0.335 |
| *p*-value | 0.187 | $p < 0.01$ | 0.022 | 0.039 | 0.039 | 0.013 |
| CI 95 percent | [-1.40,0.27] | [-1.48,-0.34] | [-1.00,-0.08] | [-0.85,-0.02] | [-0.73,-0.02] | [-0.67,-0.08] |
| Control Mean | 0.618 | 0.794 | 0.529 | 0.505 | 0.445 | 0.447 |
| Panel C. *LATE* | -2.277 | -3.676 | -2.203 | -1.822 | -1.440 | -1.400 |
| *p*-value | 0.161 | $p < 0.01$ | 0.019 | 0.032 | 0.030 | $p < 0.01$ |
| CI 95 percent | [-6.68,1.11] | [-7.36,-1.48] | [-4.83,-0.43] | [-4.03,-0.18] | [-3.37,-0.17] | [-3.04,-0.44] |
| Placebo (*p*-value) | 0.658 | 0.155 | 0.105 | 0.175 | 0.288 | 0.072 |
| Bandwidth | 55.2 | 51.4 | 55.6 | 56.1 | 62.9 | 69.7 |
| Obs(left\|right) | 986\|515 | 944\|488 | 1008\|523 | 1012\|525 | 1136\|559 | 1236\|592 |

Note: Panel A and B presents estimates of equation 1. In panel A the dependent variable is an indicator for receiving Fonden resources. In panel B the dependent variables are the annualized all-cause crude mortality rate difference for the window listed in the column title. The control mean corresponds to the left-hand-side prediction of the nonparametric regression at the threshold. Panel C reports the LATE computed as the ratio of the ITT to the first stage. The Placebo row reports the LATE *p*-value when the outcome is the AMRD computed using only pre-disaster data. Estimates in panel A and B are derived using a triangular kernel, a local linear polynomial, and the $h_{MSE}$ optimal bandwidth. The *p*-values and 95 percent confidence intervals reported are constructed using robust bias correction and clustering at the municipal level.

Next, I derive estimates of the first stage, the ITT, and the LATE. Specifically, I estimate the following equation:

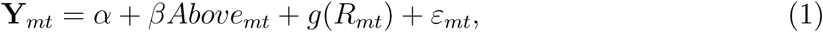

where Y_*mt*_ is the outcome variable observed in municipality *m* at disaster time *t*. When estimating the first stage the outcome is a binary variable that takes the value of 1 when a municipality receives Fonden, and 0 otherwise. When estimating the ITT the outcome is one of the AMRD. The function *g*(*R*_*mt*_) models the relationship between the outcome and the running variable *R*_*mt*_ (rainfall minus threshold). *Above*_*mt*_ is an indicator variable that takes the value of 1 when rainfall minus threshold is greater or equal than zero. In each case the parameter of interest is *β*. I also compute the ratio of the estimate of the ITT to the first stage and interpret it as the LATE under some additional assumptions.^4^

Equation 1 is estimated using non-parametric local polynomial methods, which require a choice of bandwidth selection algorithm, kernel, and local polynomial order. Unless otherwise noted, I use the bandwidth selection algorithm that minimizes the asymptotic mean squared error, from here on *h*_*MSE*_. This algorithm is optimal for point estimation (Calonico et al., 2019). Consistent with this choice, I use a triangular kernel because it provides optimal weights for the *h*_*MSE*_ (Cattaneo, Titiunik and Vazquez-Bare, 2017). Last, I follow Gelman and Imbens (2019) and use linear and quadratic local polynomials. In section 4, I show that the results are robust to the choice of these tunning parameters. In all cases, the bandwidth selection algorithm and the inference of standard errors and confidence intervals is adjusted for clustering at the municipal level.

Table 1 shows the impact of Fonden on the AMRD at four month intervals over the two year period after a disaster. The results highlight that Fonden led to a considerable and, by and large, permanent reduction in postdisaster excess mortality.

Panel A reports the first stage estimates. Consistent with the graphic depiction of the first stage the estimates reveal that being just above the threshold increases the probability of receiving Fonden by 22 to 23 percentage points relative to the municipalities just below. In all cases the coefficients are statistically significant at the one percent level.

Panel B reports the intention-to-treat estimates. Columns 1 to 6 report results from estimating equation 1 when the dependent variable is one of the AMRD computed at 4, 8, 12, 16, 20, or 24 months. The estimated coefficients highlight that Fonden led to a u-shaped reduction in postdisaster excess mortality, with the largest decrease (0.81 excess deaths per 1,000 person-years) being observed for the 8-month AMRD reported in column 2. Since the control mean (the no program counterfactual, left-hand-side prediction of the nonparametric regression at the threshold) is 0.79, this indicates that at 8-months, Fonden enabled municipalities just above the threshold to fully reduce postdisaster excess mortality relative to those just below. Moreover, as shown in columns 3 to 6, this reduction is largely permanent, as I find limited evidence of temporal death displacement (harvesting) in the two years after a disaster. To see why Fonden is not merely changing the timing of deaths, note, for example, that the 24-month AMRD includes all deaths reported between the disaster and the subsequent 24 months. If a compensating increase in fatalities followed the initial reduction observed at 8-months, I would fail to find an effect of Fonden when using the 24-month AMRD as the outcome. Instead, as shown in column 6, I find that at 24-months, Fonden led to a reduction of 0.33 excess death per 1,000 person-years, or a 75% reduction relative to the control mean.

Panel C reports estimates of the LATE. Columns 1 to 6 show that Fonden led to a reduction of between 2.2 and 3.7 excess deaths per 1,000 person-years among complier municipalities at the cut-off. Except for the coefficient for the 4-month AMRD, the estimated coefficients are all statistically different from zero at the five percent level. Next, to provide supporting evidence for the validity of the FRD design, I test whether a null effect is recovered when I use AMRD outcomes that are unaffected by Fonden because they are computed using only predisaster data. The placebo row reports the p-value when the LATE is estimated using these outcomes. Reassuringly, in all cases, I find that the coefficients are are statistically indistinguishable from zero at conventional levels. Overall, these results provide robust evidence of the capability of Fonden to reduce excess mortality in the aftermath of a disaster.

### Channels

There are four key mechanisms through which Fonden is likely to operate, to pin down the channels table 2 presents estimates of the impact of Fonden on 8-months cause-specific AMRD’s. I focus on the AMRD’s measured at 8 months because, as previously shown, the largest effect of Fonden is observed at this time. First, given that transport injuries are one of the leading causes of death and that Fonden could potentially reduce the likelihood of accidents by accelerating road reconstruction, I test whether Fonden reduces this cause of death. As shown in column 1, panels B and C, there does not appear to be an uptick in this type of death after a disaster, and I fail to find evidence supporting this channel.

**Table 2:** Impact of Fonden on cause specific AMRD’s.

|  | Transport<br>Injuries | Self-harm<br>inter-<br>personal<br>violence | Communi-<br>cable | Non-<br>commu-<br>nicable | Amenable | Amenable<br>in high<br>health<br>supply<br>areas | Amenable<br>in low<br>health<br>supply<br>areas |
| --- | --- | --- | --- | --- | --- | --- | --- |
|  | (1) | (2) | (3) | (4) | (5) | (6) | (7) |
| Panel A.<br>*First Stage* | 0.222 | 0.224 | 0.223 | 0.228 | 0.223 | 0.210 | 0.226 |
| *p*-value | $p < 0.01$ | $p < 0.01$ | $p < 0.01$ | $p < 0.01$ | $p < 0.01$ | $p < 0.01$ | $p < 0.01$ |
| CI 95 percent | [0.12,0.29] | [0.12,0.28] | [0.12,0.30] | [0.13,0.31] | [0.12,0.29] | [0.05,0.32] | [0.10,0.34] |
| Panel B.<br>*Intention*<br>*to Treat* | -0.040 | -0.040 | 0.026 | -0.464 | -0.552 | -1.050 | 0.070 |
| *p*-value | 0.215 | 0.244 | 0.322 | 0.026 | 0.013 | $p < 0.01$ | 0.805 |
| CI 95 percent | [-0.13,0.03] | [-0.11,0.03] | [-0.03,0.10] | [-0.96,-0.06] | [-1.13,-0.13] | [-2.01,-0.35] | [-0.38,0.48] |
| Control Mean | 0.028 | 0.046 | -0.031 | 0.392 | 0.532 | 0.822 | 0.120 |
| Panel C.<br>*LATE* | -0.181 | -0.177 | 0.115 | -2.032 | -2.474 | -5.010 | 0.311 |
| *p*-value | 0.188 | 0.205 | 0.299 | 0.035 | 0.015 | 0.018 | 0.799 |
| CI 95 percent | [-0.62,0.12] | [-0.52,0.11] | [-0.14,0.47] | [-4.46,-0.16] | [-5.45,-0.58] | [-11.33,-1.08] | [-1.65,2.15] |
| Placebo (*p*-value) | 0.103 | 0.424 | 0.169 | 0.146 | 0.510 | 0.365 | 0.970 |
| Bandwidth | 54.0 | 60.9 | 49.6 | 44.0 | 49.1 | 46.7 | 51.3 |
| Obs(left\|right) | 968\|513 | 1106\|550 | 923\|471 | 818\|425 | 911\|470 | 485\|244 | 426\|221 |

Note: Panel A and B presents estimates of equation 1. In panel A the dependent variable is an indicator for receiving Fonden resources. In panel B the dependent variable correspond to the cause specific 8-month AMRD listed in the column title. The control mean corresponds to the left-hand-side prediction of the nonparametric regression at the threshold. Panel C reports the LATE computed as the ratio of the ITT to the first stage. The Placebo row reports the LATE *p*-value when the outcome is the AMRD computed using only pre-disaster data. Estimates in panel A and B are derived using a triangular kernel, a local linear polynomial, and the $h_{MSE}$ optimal bandwidth. The *p*-values and 95 percent confidence intervals reported are constructed using robust bias correction and clustering at the municipal level. High and low health supply areas correspond to municipalities above and below the median of doctors per capita. In columns 6 and 7 the sample is restricted to the group listed in the column title. The *p*-value for the null hypothesis that the LATE for high and low health supply areas is equal is 0.021.

Second, I follow the literature on the impact of natural disasters on mental health (Satcher, Friel and Bell, 2007) and on conflict (Xu et al., 2016) and test whether the restoration of infrastructure and thus government presence can reduce deaths due to self-harm and interpersonal violence. As shown in column 2, I also fail to find evidence to support this mechanism.^5^

Third, given that Fonden restores access to safe water, potentially reducing deaths from waterborne or vectorborne diseases, I test whether Fonden reduces deaths caused by communicable diseases. Column 3 reports the results of this exercise. As in the previous cases, I find Fonden impact estimates that are small and statistically indistinguishable from zero. The lack of support for this channel is not surprising as the literature on epidemics following natural disasters (e.g., Watson, Gayer and Connolly, 2007) suggests that the risk of outbreaks is low unless the affected population is permanently displaced, which is unlikely in this context.

Fourth, I follow the literature on access to health services and survival from non-communicable diseases (see Di Cesare et al., 2013, and references therein) and hypothesize that by accelerating reconstruction of infrastructure (e.g., roads, electricity, safe water, and medical infrastructure), Fonden shortens disruptions to health services saving lives. Column 4 presents results from an exercise where the outcome is the 8-month AMRD for non-communicable diseases. Consistent with this mechanism, panel B shows that municipalities eligible to Fonden can fully reduce non-communicable postdisaster excess deaths relative to ineligible municipalities.

Next, in column 5, I focus on a set of conditions, sometimes referred to as amenable. That is a group of 41 conditions identified by Kruk et al. (2018) for which the risk of death can be significantly reduced by guaranteeing access to basic medical services. These conditions include, for example, rheumatic heart disease, ischemic heart disease, ischemic stroke, diabetes types 1 and 2, and chronic kidney disease. As shown in column 5 panel B, Fonden leads to a sharply estimated reduction of 0.55 amenable postdisaster excess deaths per 1,000 person-years or a 103% reduction relative to the control mean.

To further corroborate whether Fonden is operating by restoring access to existing health services, I test whether Fonden is more effective at saving lives in municipalities that initially had more extensive medical infrastructure (doctors per capita). Specifically, I split the sample at the median of doctors per 1,000 persons (0.45) and estimate the impact of Fonden in each sub-sample. Importantly, for the results’ causal interpretation, doctors per capita is measured in 2003, and as shown in figure 2 panel B, it does not change discontinuously at the threshold. Columns 6 and 7 report the results from this exercise. Panel B shows that in municipalities with above-median doctors per capita, Fonden leads to a reduction of roughly 1 amenable postdisaster excess death per 1,000 person-years. By comparison, Fonden has no impact on below-median municipalities that lack the medical infrastructure to prevent amenable deaths. Panel C provides estimates of the effects of Fonden for complier municipalities at the threshold. Using these LATE estimates, I further test and reject the null hypothesis of no differential Fonden effects (p-value 0.021). Last, note that reassuringly for all outcomes, I fail to find evidence of Fonden impact when I use placebo AMRD outcomes computed from predisaster data.

**Figure 2:**
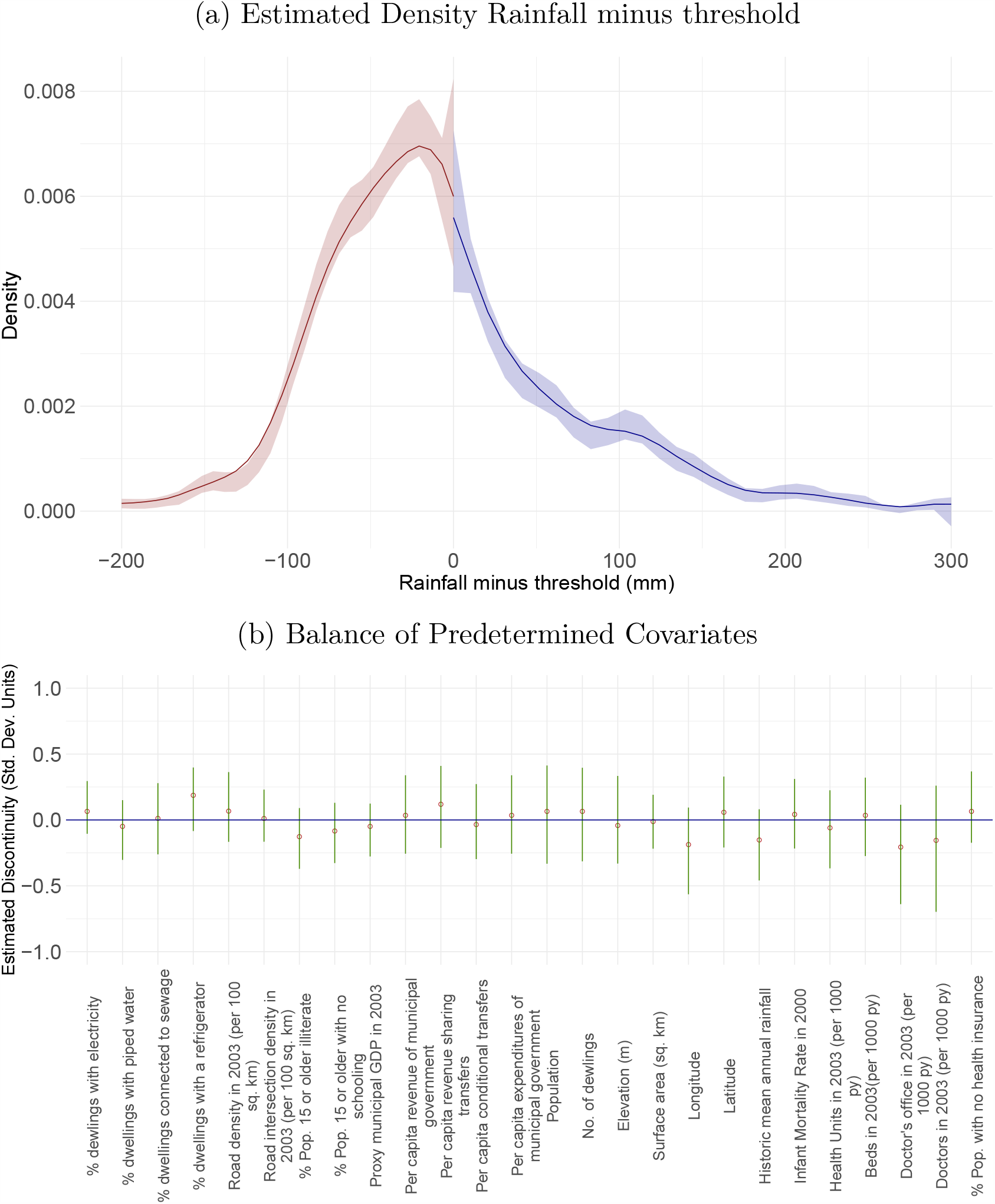
Validation and falsification of the FRD design. Notes: Panel A plots the empirical density of the running variable (rainfall minus threshold). The p-value for the null hypothesis that the density of the running variable is continuous at the threshold is 0.56. Panel B plots estimates of equation 1 using as outcome each of the variables listed. Unless otherwise stated in the label, all variables are measured in the most recent year available that predates a natural disaster used to request Fonden verification. Variables are standardized to facilitate comparison. The circles represent point estimates constructed using a triangular kernel, a local lin-ear polynomial, and a *h*_*MSE*_ optimal bandwidth. The error bars represent robust 95% confidence intervals.

In sum, this section shows that Fonden impact is concentrated in places where medical infrastructure is present and among conditions whose prognosis improves with access to these services. Accordingly, I conclude that Fonden saves lives by restoring access to medical care.

### Who’s life is saved and at what cost?

To provide a more accurate valuation of the benefits of Fonden, in this section, I study whether the program effects are heterogeneous with respect to gender and age. Specifically, I conduct exercises analogous to those of the previous sections but use sex-specific and age-specific 8-month AMRD as the dependent variables. The result are reported on Table 3. Columns 1 to 2 study the impact of Fonden by gender. The estimates reported in panel B and C reveal that Fonden reduces postdisaster excess deaths for both males and females. While the point estimates for the ITT and the LATE suggest larger reductions for males than females, given the wide confidence intervals, I cannot reject the null hypothesis of no differential gender effects (p-value 0.41).

**Table 3:** Impact of Fonden on sex and age specific AMRD’s.

|  | Male<br>(1) | Female<br>(2) | 0-14<br>(3) | 15-49<br>(4) | 50 or older<br>(5) | Life-years Lost<br>(6) |
| --- | --- | --- | --- | --- | --- | --- |
| Panel A. *First Stage* | 0.225 | 0.229 | 0.228 | 0.236 | 0.223 | 0.222 |
| *p*-value | $p < 0.01$ | $p < 0.01$ | $p < 0.01$ | $p < 0.01$ | $p < 0.01$ | $p < 0.01$ |
| CI 95 percent | [0.12,0.30] | [0.12,0.28] | [0.13,0.30] | [0.13,0.28] | [0.12,0.29] | [0.12,0.29] |
| Panel B. *Intention to Treat* | -1.055 | -0.665 | 0.172 | -0.177 | -3.376 | -36.568 |
| *p*-value | $p < 0.01$ | 0.048 | 0.264 | 0.324 | $p < 0.01$ | 0.023 |
| CI 95 percent | [-2.08,-0.34] | [-1.42,-0.01] | [-0.18,0.65] | [-0.63,0.21] | [-6.22,-1.38] | [-78.70,-5.95] |
| Control Mean | 0.865 | 0.742 | -0.169 | 0.232 | 3.357 | 39.234 |
| Panel C. *LATE* | -4.686 | -2.900 | 0.752 | -0.753 | -15.107 | -164.705 |
| *p*-value | $p < 0.01$ | 0.037 | 0.248 | 0.278 | $p < 0.01$ | 0.019 |
| CI 95 percent | [-9.97,-1.45] | [-6.71,-0.21] | [-0.74,2.88] | [-2.79,0.80] | [-30.34,-5.99] | [-376.89,-34.53] |
| Placebo (*p*-value) | 0.919 | 0.264 | 0.868 | 0.052 | 0.509 | 0.627 |
| Bandwidth | 46.4 | 63.5 | 43.7 | 70.0 | 48.7 | 52.3 |
| Obs(left\|right) | 871\|437 | 1148\|558 | 808\|423 | 1236\|591 | 901\|464 | 951\|499 |

Note: Panel A and B presents estimates of equation 1. In panel A the dependent variable is an indicator for receiving Fonden resources. In panel B columns 1 to 5 the dependent variable corresponds to the age or gender specific 8-month AMRD listed in the column title. In column 6 the outcome is the 8-month annualized life-years lost rate difference for those 50 or older. The control mean corresponds to the left-hand-side prediction of the nonparametric regression at the threshold. Panel C reports the LATE computed as the ratio of the ITT to the first stage. The Placebo row reports the LATE *p*-value when the outcome is the AMRD computed using only pre-disaster data. Estimates in panel A and B are derived using a triangular kernel, a local linear polynomial, and the $h_{MSE}$ optimal bandwidth. The *p*-values and 95 percent confidence intervals reported are constructed using robust bias correction and clustering at the municipal level. The *p*-value for the null hypothesis of no differential effects by gender is 0.41. The *p*-values for the null hypotheses of no differential Fonden effects between those 50 or older and both of the younger groups is 0.002 and 0.006.

Columns 3 to 5 study the impact of Fonden among three age groups 0-14, 15-49, and 50 or older. The results in columns 3 to 4 show no effect of Fonden among younger groups or an uptick in their deaths following a disaster. By comparison, the results in column 5 show that those 50 or older benefit disproportionately more from Fonden, with the program, entirely reducing postdisaster excess deaths among this group. As in the previous case, using the estimates for the LATE, I further test and reject the null hypotheses of no differential Fonden effects between those 50 or older and both of the younger age groups (p-values 0.002 and 0.006). The finding of Fonden effects driven by those 50 or older provides further supporting evidence for the mechanism described in the previous section, as the prevalence of non-communicable and amenable conditions is higher among this age group.

The estimates of column 5 also highlight that the reduction in annual deaths created by Fonden is of a meaningful magnitude. Among complier municipalities at the threshold, panel C shows that Fonden leads to a 15.1 per 1,000 person-years reduction in postdisaster excess deaths for those 50 or older. A simple back-of-the-envelope based on this estimate of the LATE indicates that Fonden averted up to 18,105 deaths annually.^6^

Because the benefit of the program is concentrated among older adults, I also provide a more accurate valuation of Fonden benefits by estimating the impact of Fonden on life-years lost. Specifically, in column 6, I use the 8-month annualized life-years lost rate difference for those 50 or older as the dependent variable. This outcome is computed just like the 50 or older 8-month AMRD, but the numerator is the sum of the life-years lost for this age group. That is the difference between the age-and-sex-specific counterfactual life expectancy (WHO, 2000) and the observed age at death. As can be seen in Panel C, for those 50 or older residing in complier municipalities at the threshold, Fonden leads to a reduction of 165 excess life-years lost per 1,000 person-years. An analogous back-of-the-envelope calculation based on this estimate indicates that Fonden annual benefit amounts to 197,000 life-years saved at a cost of $3,979 per life-year.^7^ This calculation greatly overstates the cost of saving lives because it assumes Fonden expenditures are meant to save lives, and not the indirect benefit of infrastructure reconstruction, whose cost is already fully offset by the accelerated economic recovery (del Valle, de Janvry and Sadoulet, 2020). Nonetheless, even under this assumption, Fonden is, by and large, cost-effective relative to other life-saving interventions that have a median cost of $63,380 per life-year saved (Tengs et al., 1995).^8^

## 4 Validation and Falsification of the FRD Design

There are two key threats to the validity of the FRD design. The first is the possibility that program thresholds may affect mortality through non-Fonden mechanisms. For example, informal use of Fonden thresholds by other government agencies for resource allocation could lead me to erroneously attribute the impact of these resources to Fonden. While I cannot rule out this possibility, an extensive review of resource allocation procedures and interviews with several senior federal and state officials failed to uncover any instance in which non-Fonden financial or in-kind transfers were allocated using Fonden thresholds. Moreover, as shown by del Valle, de Janvry and Sadoulet (2020) per capita federal transfers to local governments (including discretionary funds) do not change discontinuously at the threshold.

The second is the possibility that municipalities may try to manipulate the running variable in the hope of becoming eligible for Fonden. Gaming the Fonden verification process is difficult because the thresholds and the subset of weather stations used for verification are only known to Conagua and because rainfall information is reported nearly in real-time.

Nonetheless, to investigate whether manipulation could have taken place, I conduct three exercises. First, I follow McCrary (2008) and test whether there is a discontinuity in the running variable’s density at the threshold. In this setting, a density function that would show many more municipalities barely qualifying for Fonden (right of the threshold) than barely failing to qualify would be indicative of manipulation. Using the local polynomial density estimator of Cattaneo, Jansson and Ma (2018), figure 2 panel A plots the estimated empirical density. The figure shows both that there is no excess density just to the right of the threshold and that the running variable density is continuous at the threshold. Consistent with the figure, I cannot reject the null hypothesis that the running variable’s density is continuous at the threshold (p-value = 0.56).^9^

Second, I take advantage of the idea that if rainfall records were manipulated, the observations closest to the threshold would be those where tampering is most likely to occur. The test, therefore, consists of checking the sensitivity of the impact of Fonden (table 1 column 2) to progressively excluding observations that are within 5 mm of the threshold (2.5 mm radius). The results reported in table A2 in the appendix show that in all cases, the impact of Fonden remains statistically significant at conventional levels and that the point estimate of the LATE is of a similar magnitude.

Third, if some municipalities can game the verification process, I should observe that those just above the threshold are systematically different from those just below. To test whether municipalities’ predetermined characteristics change discontinuously at the threshold, I compiled 26 variables from the census and administrative records. These variables can be grouped into four categories. These categories include state capacity and provision of public goods, local governments’ financial capacity, geographic and demographic features of municipalities, and medical infrastructure. All variables are measured in the most recent year available that predates a natural disaster unless otherwise stated. To perform the test, I estimate equation 1 using as outcome each of the 26 predetermined covariates. Figure 2 panel B plots the resulting point estimates and 95 percent confidence intervals. The variables are measure in standard deviation units to facilitate comparison across variables. In all 26 outcomes, I fail to find evidence of a discontinuous jump at the threshold. As shown in the figure, the point estimates are small and statistically indistinguishable from zero.

On the whole, these results provide robust evidence that municipal governments were unable to sort around the threshold. The absence of manipulation provides supporting evidence both for the FRD identifying assumption and for the local smoothness assumption that underpins my interpretation of the ratio of the ITT to the first stage as Fonden’s LATE.

Next, I show that the key estimate of the impact of Fonden (table 1 column 2) is not sensitive to the choice tuning parameters used for estimation. Specifically, table A3 in the appendix provides results using alternative bandwidth selection algorithms (columns 1 to 3), degree of local polynomial (columns 4 to 5), and kernel (columns 6 to 7). In all cases, I find estimates of the first stage, the ITT, and the LATE that are of very similar magnitude and that remain statistically significant at the five-percent level.

In table A4 I further show that the estimate of Fonden impact is robust to various issues. I begin with issues related to the construction of the AMRD. Because the crude mortality rate does not account for the population’s age and sex distribution, it may not be well suited for comparisons across municipalities or over time. To address this shortcoming, I compute an 8-month AMRD that is age-sex-adjusted using as reference the age and sex distribution of Mexico in 2000. The result of this exercise is reported in column 1. Consistent with the idea that there are no systematic demographic differences among municipalities just above and just below the threshold, I find that standardization makes little difference. For example, while the estimates from table 1 column 2 indicate that Fonden fully reduces postdisaster excess deaths, the equivalent results using the standardized outcome (table A4 column 1) reveal a comparable reduction of 87%. Next, in column 2, I show that using an alternative reference pre-period for the calculation of the AMRD produces very similar results. Specifically, I compute the 8-month AMRD by subtracting the 8-month annualized mortality rate postdisaster from the average 8-month annualized mortality rate observed in the three years before a disaster. Last, while the AMRD is computed from counts of deaths that occur in the same calendar months, I further address seasonality by including calendar month fixed effects. As reported in column 3, I find that estimates are of very similar magnitudes and remain statistically significant at the one percent level. In columns 4 to 5, I test the results’ sensitivity to the exclusion of municipalities that received Fonden on consecutive years. Specifically, in column 4, I exclude municipalities that received Fonden in the year before or after a request. In column 5, I expand the window to two years before and after. In columns 6 and 7, I test whether the results are robust to excluding municipalities with extreme Fonden thresholds (bottom and top deciles of the thresholds’ distribution). Despite the smaller sample size in all cases, I find estimates that are statistically significant at the five-percent level and of a very similar magnitude.

## 5 Conclusion

Despite not experiencing more frequent or severe natural disasters than developed economies, most deaths from disasters occur in developing economies (Kahn, 2005; Strömberg, 2007). In this paper, I argue that a key feature of economic development capable of shielding the population from the worst consequences of natural disasters is the efficient provision of disaster relief. My evidence comes from studying Mexico’s Fonden program, which reduces delays and leakage of federal disaster relief by guaranteeing reconstruction resources and by enforcing the rules used for allocation and disbursement.

Taking advantage of the discontinuities created by the Fonden eligibility rules, with a fuzzy regression discontinuity design, I show that Fonden considerably reduces mortality risk from natural disasters. Specifically, I find that Fonden fully reduces postdisaster excess deaths 8-months after a disaster. This reduction is largely permanent, with a 75 percent reduction observed over the two years after a disaster. I also find that the program’s impact is driven by areas with available medical infrastructure and among conditions responsive to basic medical care. These findings suggest that Fonden saves lives by reducing disruptions in access to health services. Consistent with this mechanism, I also find that adults 50 or older benefit disproportionately from the program. Building on these findings, a simple back-of-the-envelope calculation indicates that Fonden can save a life-year at a cost of roughly $3,979 and that Fonden annually saves 197,000 life-years. This back-of-the-envelope greatly overestimates the cost of saving a life-year because this benefit occurs in addition to the Fonden led economic recovery, which is large enough to offset Fonden costs (del Valle, de Janvry and Sadoulet, 2020). Nonetheless, taking this cost figure at face value, it is still the case that Fonden ranks among the most cost-effective interventions to save a life-year.

These results are important for policymakers because they suggest that redesigning federal disaster relief programs around the idea of a pre-financed rules-based system can accelerate economic recovery and considerably reduce fatalities from natural disasters.

Two design features in this type of program require attention and could be further refined. First, the program protects taxpayer resources by conditioning payouts to the triggering of the index and the subsequent damage assessment. This setup eliminates upside basis risk (experiencing no damage and receiving a payout) but still allows for downside basis risk (experiencing damage without receiving a payout). The reduction of basis risk could be accomplished by improving the correlation between the index and damages, for example, by making complementary investments in the weather station network that provide higher resolution measurements of the hazard. Alternatively, downside basis risk could also be reduced by introducing an audit rule or a secondary index that is used in case of an appeal.

Second, the rules for the use of reconstruction resources could be further adapted to reduce deaths by incentivizing the retreat from high-risk areas and by allowing the use of resources for hardening infrastructure against hydrometeorological hazards. In this respect, Fonden’s evolution also provides a useful role model. Over time, the program adapted to allow both the use of resources to relocate infrastructure to safer areas and, under the build back better principle, to provide resources in excess of replacement costs to rebuild infrastructure at higher standards.

## Data Availability

Links for the publicly available datasets are provided in the manuscript. The assembled dataset will be available upon publication in the openICPSR repository.

## APPENDIX FOR ONLINE PUBLICATION

**Table A1:** Additional datasets and Sources.

| Variable | Source |
| --- | --- |
| *INEGI (The National Institute of Statistics and Geography)* |  |
| Population |  |
| No. of dwellings |  |
| Pop. with no health insurance |  |
| Pop. 15 or older illiterate | Census |
| Pop. 15 or older with no schooling | INEGI (2000) |
| Dwellings with electricity | INEGI (2005) |
| Dwellings with piped water | INEGI (2010) |
| Dwellings connected to sewage |  |
| Dwellings with a refrigerator |  |
| Elevation |  |
| Revenue of municipal government |  |
| Expenditures of municipal government | Public finances of municipalities |
| Total transfers | INEGI (2014) |
| Revenue sharing transfers |  |
| Conditional transfers |  |
| Municipal surface area | Municipal boundaries |
| Centroid longitude | INEGI (2013a) |
| Centroid latitude |  |
| State level GDP | INEGI (2013b) |
| *Fund for Natural Disasters Fonden* |  |
| Fonden expenditures |  |
| Fonden disbursement times | Fonden administrative record |
| Fonden planned reconstruction times | Fonden (2015) |
| *Medical infrastructure* |  |
| Health units per 1,000 person-years |  |
| Hospital beds per 1,000 person-years |  |
| Doctors offices per 1,000 person-years | DGIS (2003) |
| Doctors per 1,000 person-years |  |
| *Other Datasets* |  |
| Life expectancy by age and gender | WHO (2000) |
| Historic mean annual rainfall (20 years before Fonden) | Conagua (2015a) |
| Infant mortality rate | Conapo (2005) |
| Road intersection density (per 100 sq. km) |  |
| Road density (per 100 sq. km) | USGS (2003) |
| GDP deflator | World Bank (2010a) |
| PPP exchange rates | World Bank (2010b) |

*Note:* The terms of use of the INEGI datasets used can be found in http://en.www.inegi.org.mx/inegi/terminos.html

**Table A2:** Robustness Donut-hole analysis.

| Donut-hole<br>Radius | Intention<br>to Treat | $p$-value | CI 95% | Bandwidth | Obs | Excluded | Obs |
| --- | --- | --- | --- | --- | --- | --- | --- |
| (1) | (2) | (3) | (4) | (5) | (6) | Left<br>(7) | Right<br>(8) |
| 0.0 | -0.816 | 0.002 | [-1.48,-0.34] | 51.5 | 1432 | 0 | 0 |
| 0.5 | -0.711 | 0.006 | [-1.36,-0.23] | 55.5 | 1520 | 2 | 7 |
| 1.0 | -0.944 | 0.001 | [-1.70,-0.43] | 49.6 | 1355 | 29 | 10 |
| 1.5 | -0.830 | 0.001 | [-1.51,-0.42] | 61.8 | 1626 | 33 | 13 |
| 2.0 | -1.067 | 0.001 | [-1.89,-0.52] | 48.1 | 1291 | 40 | 21 |
| 2.5 | -0.570 | 0.057 | [-1.28,0.02] | 50.4 | 1337 | 64 | 26 |

*Note:* Robustness of the ITT estimate to the exclusion of observations that are within 2.5 mm on each side of the threshold. The dependent variable is the 8 month annualized all cause mortality rate difference. Column 1 reports the excluded radius in mm, columns 7 and 8 report the number of observations excluded at each side of the threshold. Estimates are derived using a triangular kernel, a local linear polynomial, and a $h_{MSE}$ optimal bandwidth. The p-values and 95% confidence intervals reported are constructed using robust bias correction and clustering at the municipal level.

**Table A3:**
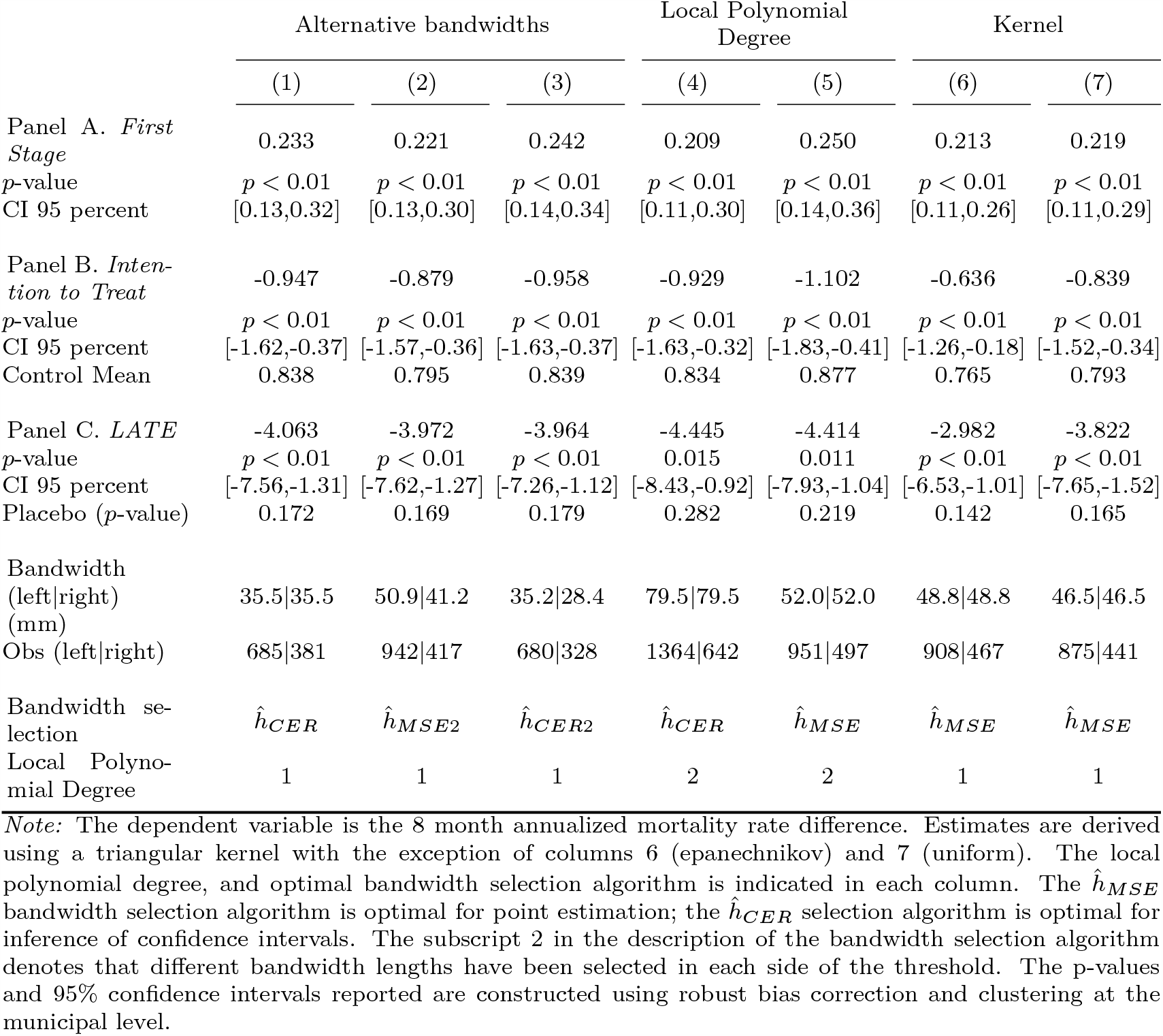
Impact of Fonden on AMRD, robustness (tuning parameters)

**Table A4:** Impact of Fonden on AMRD, robustness checks.

|  | Alternative mortality<br>rate difference<br>definitions |  | Calendar<br>Month FE | Multiple exposure<br>to Fonden |  | Extreme<br>thresholds |  |
| --- | --- | --- | --- | --- | --- | --- | --- |
|  | (1) | (2) | (3) | (4) | (5) | (6) | (7) |
| Panel A. *First Stage* | 0.222 | 0.223 | 0.236 | 0.267 | 0.289 | 0.233 | 0.201 |
| *p*-value | $p < 0.01$ | $p < 0.01$ | $p < 0.01$ | $p < 0.01$ | $p < 0.01$ | $p < 0.01$ | $p < 0.01$ |
| CI 95 percent | [0.12,0.28] | [0.12,0.29] | [0.15,0.30] | [0.15,0.36] | [0.16,0.40] | [0.12,0.29] | [0.07,0.25] |
| Panel B. *Intention to Treat* | -0.469 | -0.603 | -0.783 | -0.900 | -1.062 | -0.782 | -0.799 |
| *p*-value | 0.010 | $p < 0.01$ | $p < 0.01$ | 0.010 | $p < 0.01$ | $p < 0.01$ | $p < 0.01$ |
| CI 95 percent | [-0.90,-0.12] | [-1.14,-0.23] | [-1.39,-0.32] | [-1.77,-0.24] | [-2.00,-0.34] | [-1.46,-0.30] | [-1.53,-0.28] |
| Control Mean | 0.541 | 0.674 | 0.785 | 0.917 | 1.061 | 0.830 | 0.893 |
| Panel C. *LATE* | -2.114 | -2.703 | -3.318 | -3.370 | -3.683 | -3.356 | -3.971 |
| *p*-value | $p < 0.01$ | $p < 0.01$ | $p < 0.01$ | 0.015 | 0.011 | $p < 0.01$ | $p < 0.01$ |
| CI 95% | [-4.44,-0.62] | [-5.54,-1.06] | [-6.36,-1.32] | [-7.12,-0.78] | [-7.41,-0.95] | [-6.95,-1.38] | [-8.92,-1.72] |
| Placebo (*p*-value) | 0.100 | . | 0.223 | 0.217 | 0.247 | 0.123 | 0.075 |
| Bandwidth (mm) | 57.9 | 51.7 | 53.6 | 46.4 | 49.3 | 60.5 | 55.6 |
| Obs (left\|right) | 1047\|530 | 948\|489 | 961\|513 | 659\|314 | 497\|252 | 1020\|462 | 898\|475 |

In column 1, the dependent variable is the 8-month annualized standardized mortality rate difference. In column 2, the dependent variable is the 8-months annualized crude mortality rate difference, where the reference pre-period is the average of the 8-month annualized mortality rate observed in the 3 years before a disaster. In columns 3 to 7, the dependent variable is the 8-month annualized crude mortality rate difference. The specification in column 3 includes calendar month fixed effects. Columns 4, excludes observations with Fonden in the year before or after. Column 5, excludes observations with Fonden two years before or after. Column 6, excludes observations in the bottom decile of Fonden thresholds. Column 7, excludes observations in the top decile of Fonden thresholds. All estimates are derived using a triangular kernel, a local linear polynomial, and a $h_{MSE}$ optimal bandwidth. The p-values and 95% confidence intervals reported are constructed using robust bias correction and clustering at the municipal level.

## Footnotes

1 My weather dataset does not allow me to observe the verification process using the flooding or tropical cyclones criteria.

2 State owned assets are subject to a cost-sharing provision by which Fonden provides only partial coverage (50 percent in most cases).

3 The disaster dataset includes events up to 2012 I observe deaths up to 2017. The mortality dataset includes both the municipality of residency and the municipality where the record was created.

4 These assumptions include: monotonicity, that is, the likelihood of receiving Fonden does not decrease when rainfall is greater than the threshold; the existence of a first stage; and local smoothness (Dong, 2018). I provide supporting evidence for these assumptions in section 4.

5 I also test and find no evidence of impact when self-harm and interpersonal violence are tested separately.

6 I calculate the number of lives saved per year by restricting the sample to the municipalities that received Fonden resources and multiplying the LATE estimate for those 50 or older (column 5 panel C) by the municipal population 50 years or older. I then sum over municipalities, repeat the calculation for every year, and compute the average. This calculation assumes homogeneous treatment effects.

7 Monetary figures are in constant 2010 international dollars. The calculation procedure is analogous to the one described in the previous footnote but uses the LATE estimate in column 6 panel C. To express the figure in terms of lives saved, I multiply by minus 1. The cost per life-year is computed by dividing the life-years saved over Fonden reconstruction and administrative costs.

8 Tengs et al. (1995) report of $42,000 for the median cost is in 1993 US dollars.

9 I also use an alternative version of the test, which increases power by assuming that the cumulative density function and higher-order derivatives are the same for both groups around the threshold. I also fail to reject the null hypothesis with this test (p-value = 0.45)

## Notes

### Competing Interest Statement

The authors have declared no competing interest.

### Funding Statement

Financial support came from Georgia State University in the form of wages as an assistant professor. No external funding was received for this project.

### Author Declarations

No IRB approval was requested for this research because it is not a human subject research. The work is entirely based on publicly available data and administrative data that is not personally identifiable.

